# COVID-19 trends in Colombian regions with the highest disease burden

**DOI:** 10.1101/2020.10.09.20210187

**Authors:** Pablo Chaparro, Helmer Zapata, Isabel Hurtado, Alberto Alzate, Maria Cristina Lesmes, Sócrates Herrera

## Abstract

**Introduction:** COVID-19 pandemic is currently the most significant global public health challenge, with more than 31 million cases reported to date. Colombia first reported COVID-19 cases in the country by early March 2020, and six months later it has reached ∼750,471 clinical cases, with significant regional differences in morbidity, mortality, and hospitalization rates.

**Aims:** Identify population characteristics and hospital capacity in the 13 municipalities with the highest disease notification and examine differences in cumulative reported cases, hospitalization, and mortality rates that may explain the regional differences.

**Materials and methods:** A multi-group ecological study was performed based on the information available from public databases. Notification of cases, hospitalization, and crude mortality and age-adjusted rates were calculated.

**Results:** The municipalities with the highest COVID-1 burden at different times during the study period displayed significant differences in population density and the proportion of elderly inhabitants, indigenous and afro descendants minorities; indices of unsatisfied basic needs and multidimensional poverty index, as well as the number of hospital beds. Likewise, essential variations in notification rates, hospitalization, and mortality were observed. The highest age-adjusted of reported cases (4,219 cases) and mortality (230.4 cases) rates were found in Leticia, the lowest general hospitalization rates in Buenaventura (37.5 cases) and the lowest ICU hospitalization rates (0) in Leticia and Tumaco due to a lack of these units in these municipalities.

**Conclusion:** The probability of getting sick, hospitalized, and dying from COVID-19 appeared closely related to socio-economic, ethnic, and cultural characteristics, and also to hospital bed capacity.

## Introduction

Currently, COVID-19 pandemics represent the most significant health challenge for humanity, with ∼31 million clinical cases reported to date (Sept 22, 2020) and ∼965,000 deaths in 235 countries worldwide (1,2). The disease is produced caused by SARS-CoV-2, a coronavirus that induces a pleomorphic and broad range of clinical manifestations. Infected subjects may evolve without symptoms (asymptomatic) or develop mild, severe, and critical clinical cases frequently leading to death, particularly in elder patients with some comorbidities i.e. hypertension, cardiovascular disorders, diabetes, obesity, cancer, among other pathologies (3). The virus reached Colombia by early March 2020. A week later, the national government declared an almost full nationwide lockdown, except for essential services including the food industry and markets, pharmacies, and banks. Although the prompt lockdown significantly delayed SARS-CoV2 dissemination, fifteen days after virus entry in Colombia it caused the first COVID-19 death.

Initially, government efforts to provide early diagnosis, hospital facilities, and personal protective equipment (PPE) faced a severe global shortage of equipment, supplies, reagents, and adequate preparedness with confronting the COVID-19 challenge; therefore, a full lockdown was almost the only effective alternative. Total lockdown officially extended for about two months. As expected, its economic impact prompted the initiation on May 4 of gradual reopening, which has been associated with an immediate and progressive increase in the number of cases and deaths. As of September 18, a total of 750,471 confirmed cases and 23,850 deaths have been reported. During the first three months, 77,2% of the COVID-19 cases occurred in ten municipalities located in well-defined regions across the country: Bogotá, Cartagena, Cali, Barranquilla, Medellín, Leticia, Soledad, Villavicencio, Tumaco, and Buenaventura; however, during the last three months, Montería, Valledupar, and Bucaramanga replaced Leticia, Tumaco, and Buenaventura. In this second phase, the new top ten municipalities, each with its own geographic, demographic, social, economic, and cultural characteristics, accounted for 63.2% of the cases.

Here, we report analyses focused on the characteristics of the most affected populations and hospital beds capacity in the top ten municipalities that have reported the highest number of COVID-19 cases to date, and examine reported case rates, hospitalizations and mortality between March 6 and September 18, 2020.

## Materials and Methods

### Type of study

A multi-group ecological analysis was performed based on the information available from public databases on morbidity, mortality, and hospital bed availability.

### Geographical coverage

The thirteen study municipalities are broadly dispersed across the Colombian national geography. Barranquilla, Soledad, Monteria, Valledupar, and Cartagena are located in the northern region on the Caribbean coast, whereas Cali, Buenaventura, and Tumaco are in the western part, close or on the Pacific coast region. Bogotá, Bucaramanga, and Medellín are in the Andean zone, whereas Villavicencio is in the Eastern region, and Leticia in the southernmost town on the Amazon river (Figure 1).

**Figure 1.**
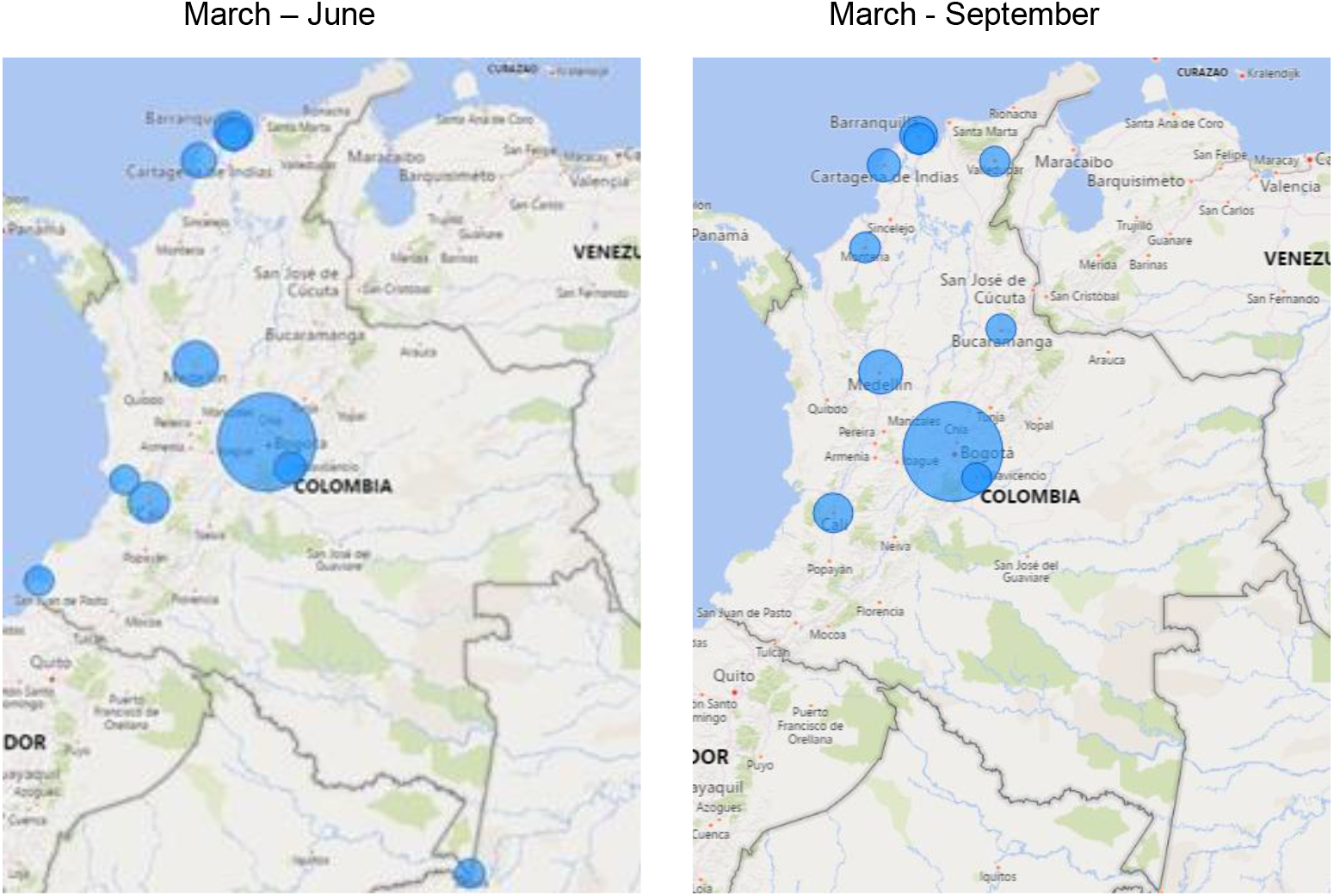
Municipalities with the highest COVID-19 burden for COVID-19. Colombia, March-September, 2020.

**Source:** Observatorio Nacional de Salud, Instituto Nacional de Salud

### Information sources

The 2018 Colombian national census conducted by the Departamento Administrativo Nacional de Estadisticas (DANE) was used to describe the characteristics of the population of each of the study municipalities (4). The indices of Unsatisfied Basic Needs (UBN) (4) and the multidimensional poverty measure (MPI) of each municipality (5) were used to determine the level of socio-economic vulnerability. The Special Registry of Health Service Providers (SRHSP; June 5 and September 18) of the Ministry of Health and Social Protection (MSP), was used to identify the capacity of hospital beds (6); and the open Data Bases of the Ministry of Information and Communications Technologies (MICT), Colombian (June 5 and September 18), were used to establish the number of people diagnosed with COVID-19, those who became ill, hospitalized and died, by municipality (7).

### Variables

municipality, total population, and population density, the average number of people per household, age median, older than 64 years, sex, ethnic self-recognition, socio-economic status, educational level, marital status, beds per 100,000 inhabitants, number of cases, dates of symptoms onset and deaths, age and medical care.

### Data analyses

The information obtained was processed in Microsoft Excel spreadsheets and analyzed with the Stata Version 12 program. COVID-19 frequency and percentage, as well as notification, mortality and hospitalization were estimated. The crude and age-adjusted rates of COVID-19 reports and mortality were analyzed; likewise, hospitalization rates were assessed. All rates were expressed per 100,000 inhabitants. For the adjustment of notification and mortality rates by age, the direct methods were considered using the World Health Organization (WHO) standard population as reference (8). Spearman correlation coefficients between mortality adjusted rate, and each of the socio-economic indicators, health providers, and hospital beds were calculated.

## Results

The Colombian population is estimated as 50,372,424 inhabitants (9), 36.4% of whom are in the municipalities analyzed between March and September. Population density varied between 9 inhabitants/km^2^ in Leticia (Amazon) and 9,926 inhabitants/km^2^ in Soledad (Caribbean). The proportion of people older than 64 years was highest in Cali (11.6%) and lowest in Leticia (4.3%). The indigenous population was highest in Leticia (42.5%), whereas the Afro-descendant population was predominant in Buenaventura (85.3%) and Tumaco (81.5%) located on the Pacific coast. Both UBN and MPI were highest in Tumaco (27.5% and 53.7%, respectively), and lowest in Bogotá (3.4% and 9.0%, respectively) (Table 1). The number of ICU beds per 100,000 inhabitants was highest in Valledupar (78.6 beds), whereas Leticia and Tumaco did not have any (0 beds) during most of the study period (Table 2). There was a significant variation in the adjusted notification of COVID-19 among the study municipalities; it was highest in Leticia (4,219 cases) and lowest in Medellín (26.6 cases) (Figure 2). The age-adjusted COVID-19 mortality rate was highest in Leticia (230.4) and lowest in Medellín (36.6) (Figure 2).

**Table 1.** Descriptive characteristics of the municipalities with the highest number of COVID-19 cases in Colombia during the period March – September, 2020.

|  | Bogotá | Medellin | Barranquilla | Cali | Cartagena | Soledad | Monteria | Valledupar | Bucaramanga | Villavicencio | Leticia | Tumaco | Buenaventura |
| --- | --- | --- | --- | --- | --- | --- | --- | --- | --- | --- | --- | --- | --- |
| Population | 7,743,955 | 2,533,424 | 1,274,250 | 2,252,616 | 1,028,736 | 665,021 | 505,334 | 532,956 | 607,428 | 545,302 | 49,737 | 257,052 | 311,827 |
| Population density | 4,360 | 6,546 | 7,676 | 4,081 | 1,840 | 9,926 | 161 | 118.6 | 3,750 | 411 | 9 | 68 | 52 |
| Average number of people per household | 2.8 | 2.9 | 3.7 | 3.0 | 3.4 | 3.7 | 3.3 | 3.6 | 3.0 | 2.96 | 3.65 | 3.01 | 3.46 |
| Median Age | 32.0 | 33.0 | 31.0 | 34.0 | 29.0 | 28.0 | 29.0 | 26.0 | 33.0 | 30.0 | 22.0 | 24.0 | 26.0 |
| Population aged 65 and over (%) | 8.9 | 10.6 | 9.7 | 11.6 | 7.9 | 6.3 | 7.6 | 6.2 | 10.5 | 7.6 | 4.3 | 6.3 | 6.4 |
| **Gender (%)** |  |  |  |  |  |  |  |  |  |  |  |  |  |
| Male | 47.8 | 47.0 | 47.9 | 46.8 | 48.1 | 48.7 | 48.51 | 48.71 | 47.5 | 49.3 | 51.5 | 48.9 | 47.4 |
| Female | 52.2 | 53.0 | 52.1 | 53.2 | 51.9 | 51.3 | 51.49 | 51.29 | 52.5 | 50.7 | 48.5 | 51.1 | 52.6 |
| **Ethnic self-recognition (%)** |  |  |  |  |  |  |  |  |  |  |  |  |  |
| Indigenous | 0.27 | 0.09 | 0.11 | 0.52 | 0.15 | 0.08 | 0.71 | 6.81 | 0.05 | 0.36 | 42.51 | 8.71 | 1.52 |
| Gypsy or Roma | 0.01 | 0.00 | 0.00 | 0.00 | 0.00 | 0.00 | 0.00 | 0.00 | 0.00 | 0.00 | 0.01 | 0.01 | 0.01 |
| Raizal | 0.01 | 0.01 | 0.02 | 0.02 | 0.05 | 0.01 | 0.01 | 0.02 | 0.01 | 0.01 | 0.03 | 0.03 | 0.02 |
| Palenquero(a) de San Basilio | 0.00 | 0.00 | 0.06 | 0.01 | 0.16 | 0.02 | 0.00 | 0.01 | 0.00 | 0.00 | 0.00 | 0.01 | 0.01 |
| Black, mulatto, Afro-descendant | 0.91 | 2.49 | 5.12 | 14.41 | 20.04 | 1.29 | 1.69 | 7.19 | 1.57 | 0.92 | 0.86 | 81.47 | 85.25 |
| Other ethnic group | 96.8 | 96.1 | 93.5 | 83.6 | 78.7 | 97.4 | 96.77 | 84.54 | 96.9 | 95.82 | 53.94 | 5.00 | 11.54 |
| Does not report | 2.0 | 1.3 | 1.2 | 1.4 | 0.9 | 1.2 | 0.82 | 1.43 | 1.4 | 2.88 | 2.66 | 4.77 | 1.66 |
| **Education level (%)** |  |  |  |  |  |  |  |  |  |  |  |  |  |
| Primary | 21.3 | 24.9 | 23.7 | 23.8 | 25.1 | 25.3 | 29.0 | 28.2 | 26.0 | 26.8 | 31.7 | 37.3 | 32.3 |
| High School | 39.6 | 44.1 | 42.9 | 44.8 | 45.9 | 48.3 | 41.5 | 40.8 | 39.4 | 41.6 | 48.2 | 44.1 | 52.8 |
| Higher | 35.0 | 27.0 | 29.7 | 27.7 | 24.9 | 23.0 | 23.5 | 24.4 | 30.5 | 25.3 | 13.1 | 5.9 | 7.9 |
| None | 1.6 | 2.4 | 2.2 | 2.0 | 2.8 | 2.3 | 4.9 | 4.9 | 2.4 | 2.7 | 3.0 | 7.5 | 4.8 |
| Does not report | 2.4 | 1.6 | 1.5 | 1.7 | 1.2 | 1.1 | 1.2 | 1.7 | 1.8 | 3.6 | 3.9 | 5.2 | 2.2 |
| **Marital status (%)** |  |  |  |  |  |  |  |  |  |  |  |  |  |
| With couple | 44.3 | 40.9 | 47.9 | 44.7 | 48.7 | 50.9 | 49.6 | 47.2 | 45.6 | 45.6 | 44.1 | 43.5 | 45.3 |
| Single | 53.8 | 57.7 | 50.8 | 53.9 | 50.2 | 48.2 | 49.6 | 51.5 | 52.9 | 51.1 | 49.9 | 51.4 | 52.7 |
| Does not report | 2.0 | 1.4 | 1.3 | 1.4 | 1.1 | 0.9 | 1.0 | 1.4 | 1.5 | 3.2 | 6.0 | 5.1 | 2.0 |
| **Socio-economic status (%)** |  |  |  |  |  |  |  |  |  |  |  |  |  |
| UBN | 3.4 | 5.2 | 9.1 | 4.1 | 12.4 | 8.0 | 18.6 | 17.7 | 5.4 | 6.3 | 27.0 | 27.5 | 16.6 |
| MPI | 9 | 12.8 | 17.4 | 11.9 | 19.9 | 18.6 | 27.1 | 30.5 | 14.2 | 15.9 | 48.4 | 53.7 | 41.0 |

**Table 2.** Availability of health services and hospital beds in Colombia, March–September, 2020.

|  | March - June |  |  |  | March - September |  |  |  |
| --- | --- | --- | --- | --- | --- | --- | --- | --- |
|  | # of health service providers | ICU beds per 100.000 inhabitants | Intermediate care unit beds per 100.000 inhabitants | Hospital beds per 100.000 inhabitants | # of health service providers | ICU beds per 100.000 inhabitants | Intermediate care unit beds per 100.000 inhabitants | Hospital beds per 100.000 inhabitants |
| Bogotá | 96 | 15.3 | 7.6 | 88.3 | 107 | 28.7 | 6.0 | 94.3 |
| Medellin | 47 | 18.2 | 12.9 | 127.1 | 41 | 30.4 | 10.7 | 130.9 |
| Barranquilla | 63 | 34.5 | 20.4 | 181.2 | 60 | 52.0 | 20.5 | 177.8 |
| Cali | 69 | 27.6 | 11.7 | 135.7 | 63 | 37.2 | 12.2 | 141.3 |
| Cartagena | 46 | 29.1 | 17.2 | 128.1 | 34 | 34.2 | 16.5 | 127.9 |
| Soledad | 15 | 5.6 | 4.5 | 42.4 | 12 | 8.7 | 7.7 | 38.2 |
| Villavicencio | 11 | 11.9 | 9.0 | 90.6 | 11 | 22.2 | 7.0 | 112.6 |
| Leticia | 2 | 0.0 | 28.1 | 168.9 | 2 |  |  |  |
| Tumaco | 3 | 0.0 | 2.3 | 50.6 | 3 |  |  |  |
| Buenaventura | 4 | 3.2 | 1.6 | 37.5 | 4 |  |  |  |
| Monteria |  |  |  |  | 27 | 70.1 | 29.7 | 222.6 |
| Valledupar |  |  |  |  | 20 | 78.6 | 24.8 | 212.8 |
| Bucaramanga |  |  |  |  | 16 | 39.2 | 14.5 | 141.3 |

**Figure 2.**
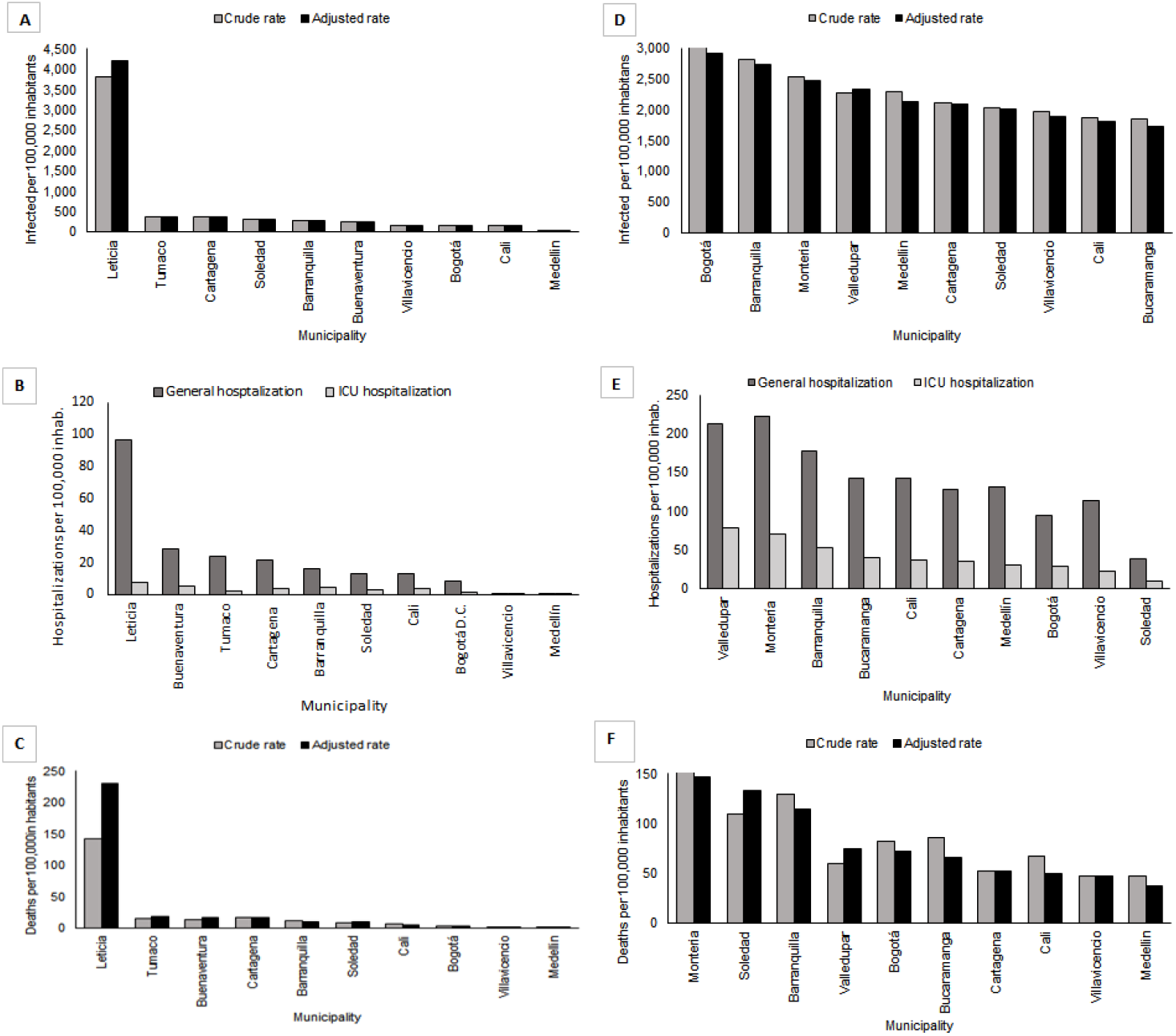
Notification of cases, hospitalizations, and deaths in municipalities with the highest COVID-19 burden between March and September 2020. COVID-19 reported cases from March-September are shown for the top 10 municipalities for March-June (**A**) and March-September (**D**); hospitalizations are shown for March-June (**B**) and March-September (**E**); and deaths for March-June (**C**) and March-September (**F**). (Data from DANE, SRHSP, MSP, MICT).

(MICT), Colombian (June 5 and September 18), were used to establish the number of people diagnosed with COVID-19, those who became ill, hospitalized and died, by municipality (7).

The Leticia, Buenaventura, and Tumaco municipalities displayed case peaks between March and June (Figure 3). In contrast, for Bogotá, Medellín, Cali, Barranquilla, Soledad, Montería, Valledupar, and Bucaramanga the peak was between June and September. In comparison, Cartagena and Villavicencio presented peaks between March and September. Spearman correlation indicated a statistical significance between the adjusted rate of mortality and case notification, population, age median age, percentage of population older than 65 years, another ethnic group (white/mestizo), education level superior and no education, UBN, MPI and number of health services providers, whereas between March and September it correlated only with having a partner (Table 3).

**Table 3.** Spearman correlation analyses.

|  | March to June | March to September |
| --- | --- | --- |
| Adjusted reporting rate | 0.867* | 0.455 |
| Population | -0.758* | -0.418 |
| Population density | -0.600 | 0.018 |
| Average people per household | 0.615 | 0.628 |
| Median Age | -0.830* | -0.579 |
| Population aged 65 and over (%) | -0.675* | -0.541 |
| Male | 0.442 | 0.383 |
| **Ethnic self-recognition** |  |  |
| Indigenous | 0.563 | 0.169 |
| Raizal of the Archipelago of San Andrés, Providencia and Santa Catalina | 0.174 | -0.174 |
| Palenquero(a) de San Basilio | 0.182 | 0.052 |
| Black, mulatto, Afro-descendant, Afro - Colombian | 0.411 | -0.103 |
| Another ethnic group (white, mestizos) | -0.782* | 0.103 |
| **Educational level (%)** |  |  |
| Primary | 0.600 | 0.248 |
| High School | 0.553 | -0.115 |
| Higher | -0.697* | -0.370 |
| None | 0.661* | -0.518 |
| With couple | 0.018 | 0.742* |
| **Economic indicators** |  |  |
| NBI | 0.891* | 0.576 |
| IPM | 0.879* | 0.552 |
| **Health service providers and hospital beds per 100,000 inhabitants** |  |  |
| # of health service providers | -0.661* | -0.091 |
| ICU beds | -0.486 | 0.358 |
| Intermediate Care Unit beds | 0.018 | 0.467 |
| General hospital beds | 0.018 | 0.298 |

\* p < 0.05

**Figure 3.**
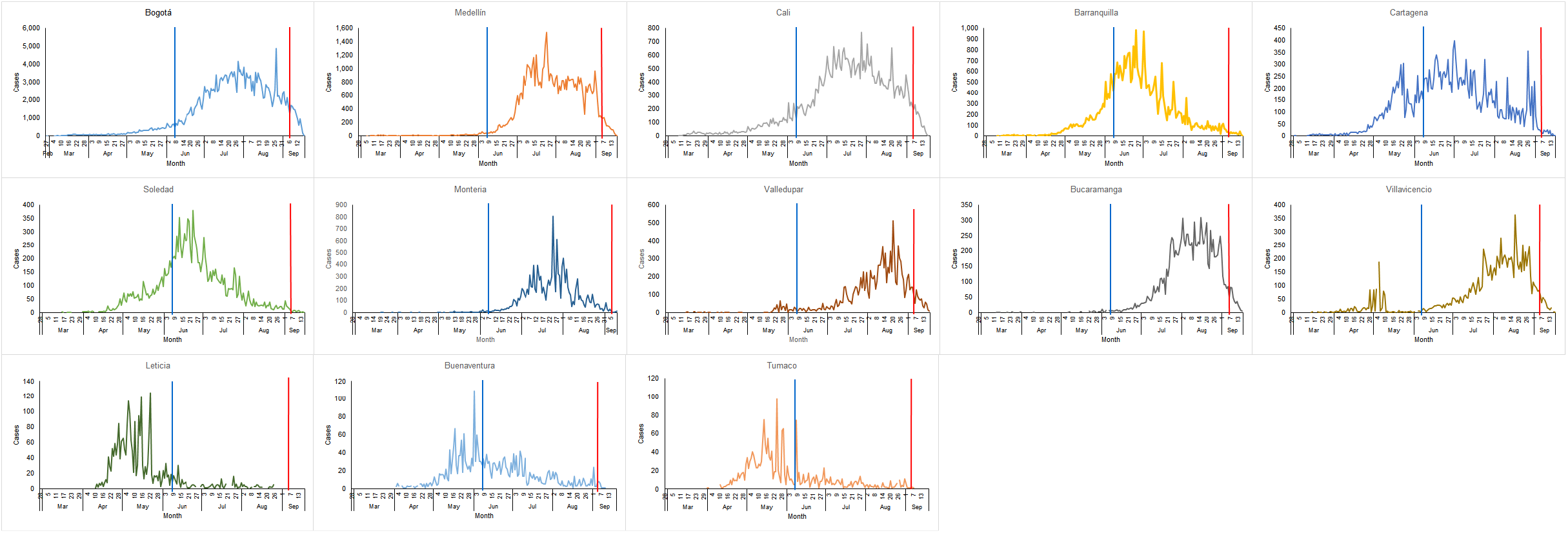
COVID-19 cases in Colombian municipalities with the highest burden March-September 2020.

## Discussion

Despite the government’s efforts and the remarkable contribution of the population to the individual recommended protective measures such as social distancing and mask usage, and the general closedown, currently Colombia is the sixth country with the highest morbidity and eleventh in mortality attributed to COVID-19 worldwide (1). Regardless of severity, during a pandemic the burden on health services is great and exposes any weakness of the system (10). Furthermore, maintaining sustainable critical care services is a crucial consideration for the health system (11). The analyses reported in this study revealed significant variations in the notification of both general and ICU hospitalizations and mortality rates in the most affected municipalities during the six months study period. Whereas Leticia, with the highest proportion of the indigenous population with the highest MPI, displayed one of the highest rates of COVID-19 hospitalization and death rates, Medellín showed the lowest notification and mortality rates. These differences correlate with the socio-economic conditions of both municipalities. In Leticia, a large part of the indigenous population has a high degree of socio-economic marginalization, living in distant and isolated areas of the Amazon forest region, and communities are exposed to several factors that endanger their health status (12). In this region, search for medical care demands traveling long distances, usually using primitive, inefficient, and expensive transportation means, which are consequently time-consuming, notably affecting subsistence and loss of work. These hurdles result in increased time between symptoms onset and adequate medical care, which is usually provided in primitive health service conditions (13). Although the isolation of these communities from the main population center of Colombia likely delayed the arrival of COVID-19, its proximity to Tabatinga, the closest town on the Brazilian border, and Santa Rosa de Yavari on the Peruvian border, might have accelerated the disease (virus) arrival to this isolated region where health infrastructure was not prepared for the pandemic. Second, people were skeptical about the real danger of the disease (14).

When comparing the municipalities of the Caribbean (Cartagena, Barranquilla and Soledad) and the Pacific (Buenaventura and Tumaco) coasts, remarkable differences were also observed. While the Caribbean has a high population density, better socio-economic conditions, and better hospital infrastructure, the Pacific region is populated by Afro-descendant minorities, traditionally affected by poverty (15) with poor housing infrastructure, malnutrition and limited access to health care (16,17). Afro-descendant communities are predominant in the Pacific regions. It is also possible that the difference due to ethnic self-recognition is attributable to the access that this population may have to care in health services (18); however, the concept of ethnicity is linked to its own historical, political, and social baggage (19). In general, biological susceptibility, geographic, socio-economic, and cultural factors, along with pre-existing health conditions and access to medical care, can help explain the higher rates of infection and death in people of African descent (19,20). On the other hand, indigenous populations are predominant in the Amazon and Pacific regions. Both minorities displayed significantly more cases than mestizo populations predominant in the rest of the country.

The differences in morbidity and mortality due to COVID-19, associated with economic and social conditions, have been described in racial and ethnic North American minorities (21). Social, economic, and cultural differences reported here are similar to those presented by Wadhera et al., who found that ethnic minorities living in poverty in New York, who have a low educational level, had higher COVID-19 hospitalization and related mortality (22). Furthermore, in Colombia another study that evaluated the influence of poverty and inequity on COVID-19 found that the income losses induced by the closedown affected significantly more the vulnerable sectors (23).

Regarding the provision of health services, there was a great variation in those of general hospitalization and ICU hospitalization in the 13 municipalities. Much of the variation could be related to NBI and MPI levels. This variation in the provision of health services occurred both within and between municipalities. In the last month efforts have been made to minimize these variations aimed at guaranteeing adequate access for the entire population and avoiding the excessive use of potentially expensive resources.

The results presented here should be interpreted with caution. These are partial results prior to September 18, 2020; these trends may be currently changing according to the evolution of the disease in the population. As for hospital and ICU services, this study was an attempt to measure their supply. It is possible that the municipalities where the greatest burden of the disease was reported are not the most representative of the socio-economic conditions of the regions to which they belong. In conclusion, the probability of getting sick, hospitalization, and death from COVID-19 between municipalities may be related to socio-economic characteristics and the local capacity of hospital beds. Populations with worse conditions bear the greatest burden of disease. The results presented can contribute to data-based decision-making related to the availability of health services affected by morbidity and mortality due to COVID-19 and, in particular, the optimal capacity of hospital services and ICUs.

## Data Availability

the data mentioned in the manuscript are available
1. The 2018 Colombian national census conducted by the National Administrative Department of Statistics (DANE)

https://www.dane.gov.co/index.php/estadisticas-por-tema/demografia-y-poblacion/censo-nacional-de-poblacion-y-vivenda-2018

https://www.dane.gov.co/index.php/estadisticas-por-tema/demografia-y-poblacion/censo-nacional-de-poblacion-y-vivenda-2018

https://www.dane.gov.co/index.php/estadisticas-por-tema/pobreza-y-condiciones-de-vida/pobreza-y-desigualdad/medida-de-pobreza-multidimensional-de-fuente-censal

https://prestadores.minsalud.gov.co/habilitacion/

https://minsalud.maps.arcgis.com/apps/opsdashboard/index.html#/1de89936b24449edb77e162d485ed5d9

https://www.datos.gov.co/Salud-y-Protecci-n-Social/Casos-positivos-de-COVID-19-en-Colombia/gt2j-8ykr

## Conflict of interest statement

The authors declare no conflicts of interest.

## Financing

This work was carried out with operating resources from the National Health Observatory of the National Health Institute, Gobernación del Valle and Caucaseco SRC.

## Notes

### Competing Interest Statement

The authors have declared no competing interest.

### Funding Statement

Neither the authors or the institutions belonging to the study have received payments or services from a third party for any aspect of the submitted work.
No external financing was received

### Author Declarations

It is an ecological study based on secondary sources open to the public

